# Changes in Child Suicide Rates and Characteristics During the COVID-19 pandemic in England

**DOI:** 10.1101/2025.07.07.25330625

**Authors:** David Odd, Duleeka Knipe, Tom Williams, Sylvia Stoianova, Prathiba Chitsabesan, Karen Luyt

**Author notes:** Corresponding author Prof Karen Luyt.

## Abstract

**INTRODUCTION:** Since the start of the COVID-19 pandemic inequalities around child mortality are likely to have increased. Suicide in young people has risen in many countries over the last 10 years, and suicide in particular may have been expected to increase over the course of the lockdown, as rates of mental health needs increased.

**AIM:** The aim of this work was to report any changes, and characteristics of children dying of suicide in England, before, and during the COVID pandemic.

**METHODS:** Child deaths from suicide, reported to the National Child Mortality Database, occurring between 1st April 2019 and 31^st^ March 2023 were identified, and linked to demographic data, death-review data and routine Hospital Episodes Statistics (HES) data (preceding the death). Routine HES data was used to identify mental health disorders and self-harm events. Temporal trends across the time period were quantified, alongside any changes in sociodemographic characteristics. Using Case-Cross Over methodology, we investigated the relative risk of suicide, after recent HES-coded events.

**RESULTS:** In total there were 498 deaths likely due to suicide, during the 4 year period. Overall risk of death by suicide was 14.31 (13.08-15.63) per 1,000,000 CYP per year. Overall, there was little evidence that risk (p=0.863) or method (p=0.199) changed over the period (p=0.863). There was evidence that the relationship between deprivation and suicide risk was different between ethnic groups (both p<0.001), with decreasing deprivation associated with increasing risk of suicide in white children (IRR 1.12 (1.03-1.21)), and decreasing risk in Asian (IRR 0.52 (0.41-0.65)), Black (IRR 0.31 (0.21-0.44)) and Mixed/Other ethnicity (IRR 0.73 (0.60-0.89) children. Only a recorded diagnosis of self-harm was more common before the death than in the preceding control periods (OR 8.99 (4.27-18.94)).

**CONCLUSION:** In England, suicide rates do not appear to be increasing, and the methods of suicide remain static. However, the role of deprivation and suicide risk appears to be different between children of different ethnic groups, and while hospital admission and a recorded diagnosis of mental health disorder does not appear to predict suicide in the subsequent month, there was a strong association with self-harm events.

## INTRODUCTION

Since the start of the COVID-19 pandemic there have likely been over 14 million excess deaths worldwide(1), although during the pandemic the number of deaths from COVID in children and young people (CYP) was low(2), with overall mortality for CYP reducing over, and around, the period of lockdown(3, 4). Rates of probable mental health disorders have increased in CYP following the pandemic. The rate of probable MH disorder for 8–16-year-olds has increased from 12.5% to 20.3% between 2017 and 2023, and for 17–19-year-olds, it has increased from 10.1% to 23.3% over the same period (5). A previous NCMD report found that 55% of CYP who had died by suicide had a mental health need, 49% a history of risk taking behaviours including self-harm, 16% neurodevelopmental conditions and 9% concerns about sexual orientation, sexual or gender identity (6).

Despite this increase, global evidence has shown that there has been little impact of the pandemic on suicide rates in CYP, and limited evidence that rates have changed in CYP in England during the early part of the pandemic(7, 8) (9)

However, inequalities around child mortality are likely to have increased since the start of the pandemic (10). Determinants of mental ill health interact with inequalities in society, placing some people at higher risk of poor mental health than others or poorer access, experience and outcomes from services including those living in areas of greater deprivation, from ethnic minority backgrounds or those who identify as LGBT plus(11). The current evidence on the impact of the pandemic on CYP suicide in different marginalised groups is scarce(12). In England and Wales the rate of suicide in female CYP has been steadily increasing (ONS) and recent evidence indicate ethnic differences in the rates of suicide (13, 14). Self-harm rates in England have also been increasing over time, with evidence that this increase is across all ethnic groups (15). Previous research, in England and Wales, has indicated that the rate of suicide in CYP is higher in some ethnic groups than others, with individuals from a Mixed and White heritage background having the highest rates(13). Additionally, suicide is more common in more deprived areas in England(16) but this finding appears to be limited to adults, as previous findings in CYP suggest limited social gradient(7) . Evidence during the pandemic suggested that individuals from certain ethnic minority backgrounds who live in more economically deprived areas experienced higher COVID-19 mortality than their White counterparts(17). It is, therefore, possible that whilst we do not see a difference by deprivation in CYP suicide mortality as a whole, the interaction between ethnicity and deprivation might alter this association.

The prevalence of psychiatric morbidity in individuals who die by suicide is high, while only a third of young people who died by suicide were in contact with mental health services at the time(6). Accessing support for mental ill health including self-harm prior to suicide provides an opportunity for intervention.

The aim of this work is to explore whether the sociodemographic profile of CYP who died by suicide changed between 2019 and 2023 and whether, for those individuals who accessed hospital care, the type of mental health contact prior to death differed between the time points and overall. Additionally, we explored whether the associations between sex, age, deprivation, and region with suicide differed by ethnicity.

## MATERIALS AND METHODS

### NCMD Methods

The NCMD collects data from the 58 Child Death Overview Panels (CDOPs) that review the deaths of all children who die before their 18^th^ birthday in England. There is a legal responsibility for CDOPs to notify NCMD of any death of someone aged under 18 years within 48 hours of it occurring, using an electronic system. The NCMD commenced data collection on 1^st^ April 2019.

### Inclusion criteria

Child deaths occurring between 1st April 2019 and 31^st^ March 2023 that were categorised as suspected suicide at the point of notification (48 hours) by one or more of the NCMD clinical team, or where the CDOP had reviewed the death and assigned the primary category ’Suicide or deliberate self-inflicted harm’ (Category 2) were extracted. Information assigned at review stage was used in precedence over that assigned at notification. Deaths that were initially coded as suspected suicide which went on to be reviewed by CDOP as any other primary category were excluded. In addition, deaths that were reviewed as ‘Death as the result of substance misuse (excluding deaths as a result of a deliberate overdose)’ within category 2, or where all 3 clinicians coded ’substance misuse’ were excluded.

Deaths that were reviewed by CDOP as sub-category ’Suicide (where the panel feels the intention of the child was to take their own life)’ or where all 3 clinicians coded ’suicide’ were included for analysis. For remaining disagreements (i.e. those where the sub-category at review was not known, or where the review was ongoing and suicide was coded 1 or 2 times), all information within NCMD was reviewed and validated by an expert clinical researcher. For this, the same methodology was followed as used in our previous studies. The probability that each death was intentional suicide will be categorised using the following categories: High, Moderate, Low or Unclear. Deaths that were assessed as High or Moderate were included as likely suicides.

### Coding Methods

Information recorded within the child death notification form for deaths occurring between 1^st^ April 2019 and 31^st^ March 2023 were extracted, including:

- Sex of individual (female, male, other (including not known))
- Ethnic Group (Asian or Asian British, Black or Black British, Mixed, Other, Unknown, White)
- Age at death
- Deprivation measures of the child’s home address, derived from the Index of Multiple Deprivation (IMD). For this work each child was placed in the quintile of deprivation, with 1 being the most deprived, and 5 the least(18).
- Region of England of the Child Death Overview Panel (CDOP) responsible for the child death review

Additionally, all information available within the NCMD record at the time of data extraction (finalised January 2024) was reviewed to identify:

- Method of death
- Deaths where the child identified as LGBT+. This included all children whose sexual orientation was reported as ‘bisexual’, ‘gay’, ‘lesbian’ or ‘other’ and also where the case information recorded that the child was trans (used as the umbrella term for transgender, gender questioning, gender transitioning, or gender dysphoria)(19).

### Hospital Episodes Statistics data

For deaths between 1^st^ April 2019 and 31^st^ March 2022, Hospital Episode Statistic (HES) admitted patient care data was requested from acute (non-mental health) hospital admissions. Data was received which contained all ICD-10 diagnosis codes from each episode of acute inpatient care the child had since birth (including evidence of self-harm). ICD-10 diagnosis codes (in any position) recorded within the HES data were linked to groups of mental health and neurodevelopmental disorders by ICD codes, where ’*’ denotes any character:

- Previous Evidence of Self Harm: X6* and X7*
- Attention Deficit Hyperactivities Disorders: F900
- Autistic Spectrum Disorders: F840, F841, F845, F848 ,F849
- Eating Disorders: F50*
- Anxiety Disorders: F41*
- Post-Traumatic Stress Disorder: F431
- Mood Disorders: F3*
- Schizophrenia related disorders: F20*, F22*, F23*
- Any mental health disorder: Any of the above codes.

### Statistical Analysis

Initially we compared the number of children dying of likely suicide across the time period, split by year (April to March). Sociodemographic characteristics of cases across the time period were compared using Fishers exact test for categorical data and Mann-Whitney U for age and deprivation quintile.

We then repeated this comparison looking at characteristics identified from the child death review information (LGBT+, method of death). Next, we identified the number of children linked to HES data, and the prevalence of these conditions across the first 3 years of the time frame. Next, using Case-Cross Over methodology(20), we investigated the relative risk of likely suicide, after recent HES – ICD 10 coded events. All CYP data was interrogated for ICD-10 coded events (as above) occurring between 2 and 28 days before the death (i.e. at least 48 hours before the death). For comparison, 4 other ‘control’ days were identified, moving back in time by 4 weeks steps (i.e. 28 days before the death, 56 days before the death etc), and ICD-10 coded events identified in a comparable time period before them. The presence, or not, of an event was initially compared using a univariable condition logistic regression, comparing the odds, for each child, of an event in the period prior to death, with periods prior to this (to derive an Odds Ratio (OR)). This model was repeated, using a random-effect model, with the CYP’s ID as the grouping variable, and then subsequently, adjusting for year of death (coded as above) and age at the event date as a linear term. Next, we derived the risk of death overall, and by demographics of the child. Underlying population structure was derived from the Office of National Statistic 2021 UK census(21) for children between 5 and 18 years. Risk and confidence intervals were derived assuming a Poisson distribution and relative rates (defined as Incidence Rate Ratios (IRR)) were derived with counts of death collapsed to numbers per month, and using a random effects model clustered by month of death to reduce the effect of seasonal variation. To identify the relative rate of dying across the characteristics derived a p-value was obtained by comparing a model with, or without, the measure of interest (e.g. sex) to test if the value appeared to be different by the measures (tested using the likelihood ratio test). Next, we derived the relative rate of death by child characteristics across the time frame, with April 2019 to March 2020 being the reference year, and a test for linear trend being performed across the entire time period.

Finally, using a Poisson regression model we derived the relative rate of death by child characteristics across the time frame, split by the ethnic group of the child. Due to smaller numbers, groups “Mixed” and “Other” were combined. Deprivation was used as an ordinal measure, to assess the relative rate of likely suicide as deprivation reduced in each ethnic groups, alongside the risk of likely suicide per 1,000,000 person years (5-17 year olds) for each quintile of deprivation. The model we repeated, as above, adjusting for the other available covariates.

Analysis was performed using Stata Version 17. All tests were two-sided. No *a-priori* level of statistical significance was defined(22).

## RESULTS

In total there were 498 deaths, identified as due to likely suicide or self-harm, during the 4 year period (Table 1). There was little evidence that the profile of sex (p=0.496), ethnicity (p=0.638), age category (p=0.313) or region of England (p=0.627) varied over the 4 years. There was however variation in the deprivation level seen across the period (p=0.026), although a clear linear pattern was not apparent.

**Table 1.**
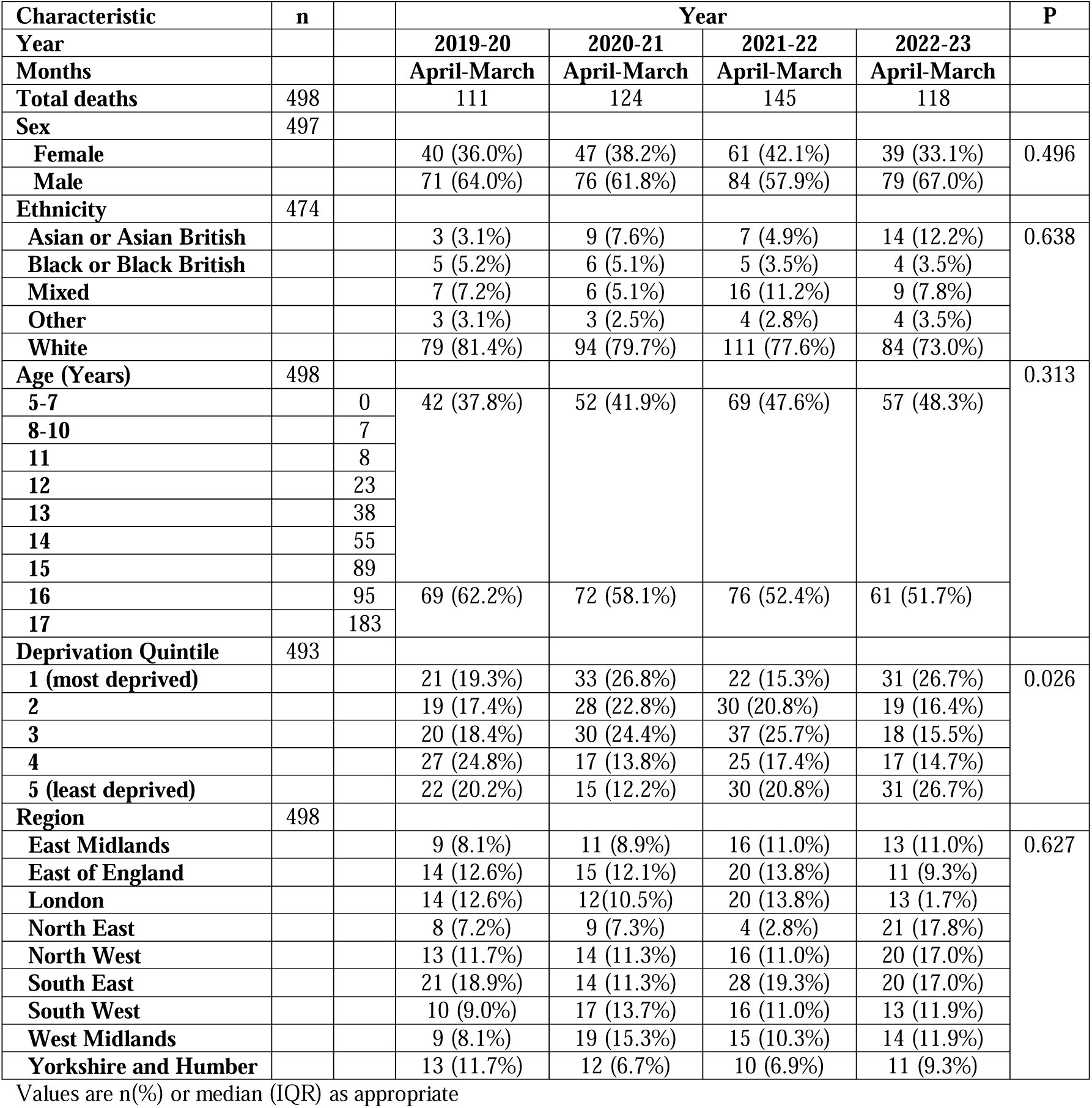
Changes in characteristics of childhood likely suicide deaths by year (starting April 1^st^)

The number of children identified as LGBT+ across the 4 years appeared to vary (p=0.027), with the highest number in 2021-22 (27 (18.2%)); although there was little evidence of a trend (p_trend_=0.4409) (Table 2). A similar profile was seen for trans CYP (p=0.045), with the highest number in 2021-22 (15 (10.3%)) compared to other years, but little evidence of an overall trend (p_trend_=0.1980). Method of death did not appear to differ across the 4 years. Within the group where poisoning was identified as the mode of death, no clear common agent, or temporal trends were identified (data suppressed due to small numbers throughout). Where a location was identified in the reported data (n=39), most events occurred in the home (32 (82.1%)), with 4 (10.3%) occurring in a public place, or place of education.

**Table 2.** Additional Data from Child Death Overview Panels (CDOP) and Health Episode Statistics (HES) linkage.

| Characteristic | Total |  | Year |  |  |  | P | Ptrend |
| --- | --- | --- | --- | --- | --- | --- | --- | --- |
|  |  |  | 2019-20 | 2020-21 | 2021-22 | 2022-23 |  |  |
|  |  |  | April-March | April-March | April-March | April-March |  |  |
| All deaths | 498 |  | 111 | 124 | 145 | 118 |  |  |
|  | CDOP data |  |  |  |  |  |  |  |
| LGBT+ | 59 |  | 11 (9.9%) | 10 (8.1%) | 27 (18.6%) | 11 (9.3%) | 0.027 | 0.4409 |
| Not trans | 30 |  | 7 (6.3%) | 6 (4.8%) | 12 (8.3%) | 5 (4.2%) | 0.515 | 0.8257 |
| Trans | 29 |  | 4 (3.6%) | 4 (3.2%) | 15 (10.3%) | 6 (5.1%) | 0.045 | 0.1980 |
| Method | 498 |  |  |  |  |  | 0.199 |  |
| Fall or fracture |  |  | 10 (10.4%) | 13 (10.5%) | 13 (9.0%) | 5 (4.2%) |  |  |
| Firearms |  |  | NA | NA | NA | NA |  |  |
| Hanging or Strangulation |  |  | 73 (75.8%) | 93 (75.0%) | 96 (66.2%) | 82 (69.5%) |  |  |
| Jumping or lying in front of a moving object |  |  | 16 (14.4%) | 12 (9.7%) | 18 (12.4%) | 17 (14.4%) |  |  |
| Other |  |  | NA | NA | NA | NA |  |  |
| Poisoning |  |  | 7 (6.3%) | 6 (4.8%) | 11 (7.6%) | 8 (6.8%) |  |  |
| Unclear |  |  | NA | NA | NA | NA |  |  |
|  | HES data |  |  |  |  |  |  |  |
| Deaths Linked | 275 |  | 94 | 101 | 80 | NA | NA | NA |
| Self-Harm | 44 |  | 15 (16.0%) | 13 (12.9%) | 16 (20.0%) | NA | 0.430 | 0.5028 |
| ADHD | 8 |  | NA | NA | NA | NA | 0.963 | 0.7891 |
| ASD | 14 |  | 7 (7.5%) | 3 (3.0%) | 4 (5.0%) | NA | 0.364 | 0.4301 |
| Eating Disorder | 13 |  | 4 (4.3%) | 4 (4.0%) | 5 (6.3%) | NA | 0.744 | 0.5527 |
| Anxiety Disorder | 22 |  | 6 (6.4%) | 8(7.9%) | 8 (10.0%) | NA | 0.681 | 0.3832 |
| PTSD | 1 |  | NA | NA | NA | NA | 0.380 | 0.2318 |
| Mood Disorder | 24 |  | 7 (7.5%) | 8 (7.9%) | 9 (11.3%) | NA | 0.633 | 0.3867 |
| Schizophrenia | 2 |  | NA | NA | NA | NA | 0.176 | 0.9276 |
| Any of above | 61 |  | 18 (19.2%) | 21 (20.8%) | 22 (27.5%) | NA | 0.382 | 0.1947 |

Values are n(%)
NA – Not Available

There was little to suggest a difference in underlying psychiatric morbidity across the 4 years for any a-priori proposed condition (all p>0.10). In total 61/275 (22.2%) of CYP who died by likely suicide had an identified acute-hospital recorded mental health diagnosis. In addition, of those trans CYP, 5 (33.3%) had a previous acute-healthcare admission and an ICD 10 coded recording of a psychiatric disorder, and 2 (13.3%) had ASD.

There was little evidence (limited by the small numbers), that eating disorder, PTSD, mood disorder or schizophrenia mental health diagnosis recorded during acute hospital admission, were more common before the death, than in the preceding control periods (Table 3). However, children were far more likely to have an ICD-10 recorded diagnosis of self-harm (Conditional Regression OR 9.30 (4.27-20.26); Adjusted Random Effects OR 8.99 (4.27-18.94)) during an acute-healthcare contact in the period immediately before their death, than in the preceding weeks. There was also weak evidence that ICD 10 coded anxiety events were more likely (Conditional Regression OR 4.00 (1.00- 6.00)); Adjusted Random Effects OR 3.89 (0.96-15.83)).

**Table 3.** Case Cross-over Analysis for Health Episode Statistics (HES) coded events in the immediate period before death, compared to previous periods.

| Characteristic | Total | Odds Ratio (95% confidence interval) |  |  |
| --- | --- | --- | --- | --- |
|  |  | Conditional Regression | Random Effects (Unadjusted) | Random Effect (Adjusted)* |
| **Deaths Linked** | 275 |  |  |  |
| **Self-Harm** | 44 | 9.30 (4.27-20.26) | 8.68 (4.15-18.15) | 8.99 (4.27-18.94) |
| **Eating Disorder** | 13 | 0.54 (0.06-4.77) | 0.57 (0.07-4.65) | 0.59 (0.07-4.834) |
| **Anxiety Disorder** | 22 | 4.00 (1.00-6.00) | 4.05 (1.01-16.30) | 3.89 (0.96-15.83) |
| **PTSD** | 1 | NA | NA | NA |
| **Mood Disorder** | 24 | 3.09 (0.58-16.42) | 2.42 (0.57-10.18) | 2.14 (0.50-9.17) |
| **Schizophrenia** | 2 | NA | NA | NA |
| **Any of above** | 61 | 6.87 (3.75-12.59) | 4.63 (2.82-7.59) | 4.70 (2.85-7.76) |

Values are OR (95% CI)
NA – Not Available

Overall risk of death by likely suicide was 14.31 (13.08-15.63) per 1,000,000 CYP per year over the 4 years (Table 4). When comparing risk between demographic categories, males had a higher risk than females (IRR 1.58 (1.31-1.89), p<0.001)), as did CYP aged 16-17 compared to those 5-15 (IRR 7.25 (6.07-8.65), p<0.001). Both associations persisted in the adjusted analysis (Males vs Females IRR 1.59 (1.32-1.92)), CYP 16-17 vs 5-15 years IRR 7.02 (5.85-8.42)). There was also strong evidence that risk varied by ethnicity, with the highest risks seen in CYP from white (14.62 (13.17- 16.19) per 1,000,000 children per year), Other (15.08 (8.25-25.31 per 1,000,000 children per year) or Mixed (16.61 (11.70-22.90) per 1,000,000 children per year) backgrounds. In the unadjusted, and adjusted analysis, CYP from Asian or Asian British Backgrounds had lower risks than those from White backgrounds (IRR 0.59 (0.41-0.86)). There was weak evidence (p=0.0458) that the risk of death by likely suicide varied by measures of local deprivation, with the lowest risk seen in the least deprived areas (IRR 0.73 (0.55-0.96)). Associations attenuated with adjustment for other demographic factors (e.g. least deprived quintile vs most deprived IRR 0.84 (0.62-1.13). There was little to suggest an overall difference in the risk between different regions of England (p=0.4248), although when compared to the reference area of London, two regions appeared to have higher risks in the unadjusted analysis (East Midlands IRR 1.52 (1.04-2.21), South West 1.57 (1.09-2.26). However, both these associations attenuated after adjustment for other demographic factors.

**Table 4.** Absolute, and relative risks of likely suicide by child characteristics, and over time.

| Category | n | Rate per 1,000,000 per year* | P | Incidence Rates Ratios (between groups) |  | Incidence Rate Ratio (IRR) per year |  |  |
| --- | --- | --- | --- | --- | --- | --- | --- | --- |
|  |  |  |  | Unadjusted | Adjusted | IRR | P<sub>trend</sub> | P<sub>interaction</sub> |
| **All Suicides** | 498 | 14.31 (13.08-15.63) | - | - | - | 1.03 (0.96-1.11) | 0.434 | - |
| **Sex** | 497 |  | <0.001 |  |  |  |  | 0.863 |
| Female |  | 11.03 (9.51-12.73) |  | Ref | Ref | 1.02 (0.90-1.16) | 0.726 |  |
| Male |  | 17.38 (15.50-19.43) |  | 1.58 (1.31-1.89) | 1.59 (1.32-1.92) | 1.04 (0.94-1.14) | 0.463 |  |
| **Ethnicity** | 474 |  | 0.0008 |  |  |  |  | 0.174 |
| Asian or Asian British |  | 7.72 (5.31-10.84) |  | 0.53 (0.37-0.75) | 0.59 (0.41-0.86) | 1.48 (1.08-2.02) | 0.015 |  |
| Black or Black British |  | 9.74 (5.95-15.04) |  | 0.67 (0.42-1.05) | 0.75 (0.47-1.21) | 0.87 (0.60-1.28) | 0.489 |  |
| Mixed |  | 16.61 (11.70-22.90) |  | 1.14 (0.81-1.59) | 1.31 (0.93-1.85) | 1.11 (0.84-1.47) | 0.454 |  |
| Other |  | 15.08 (8.25-25.31) |  | 1.03 (0.61-1.76) | 1.17 (0.68-2.02) | 0.89 (0.57-1.39) | 0.622 |  |
| White |  | 14.62 (13.17-16.19) |  | Ref | Ref | 1.04 (0.95-1.14) | 0.392 |  |
| **Age (Years)** | 498 |  | <0.001 |  |  |  |  | 0.106 |
| 5-15 years |  | 7.43 (6.48-8.48) |  | Ref | Ref | 1.11 (0.99-1.24) | 0.084 |  |
| 16-17 years |  | 53.82 (47.68-60.53) |  | 7.25 (6.07-8.65) | 7.02 (5.85-8.42) | 0.97 (0.88-1.08) | 0.625 |  |
| **Deprivation Quintile** | 493 |  | 0.0458 |  |  |  |  | 0.437 |
| 1 (most deprived) |  | 16.18 (13.26-19.55) |  | Ref | Ref | 1.04 (0.88-1.23) | 0.623 |  |
| 2 |  | 15.19 (12.31-18.52) |  | 0.94 (0.71-1.24) | 0.92 (0.69-1.22) | 0.98 (0.82-1.16) | 0.812 |  |
| 3 |  | 16.29 (13.32-19.72) |  | 1.01 (0.77-1.32) | 1.02 (0.77-1.35) | 1.00 (0.84-1.18) | 0.963 |  |
| 4 |  | 12.26 (9.81-15.15) |  | 0.76 (0.57-1.01) | 0.82 (0.61-1.10) | 0.95 (0.79-1.14) | 0.554 |  |
| 5 (least deprived) |  | 11.76 (9.55-14.34) |  | 0.73 (0.55-0.96) | 0.84 (0.62-1.13) | 1.20 (1.01-1.43) | 0.041 |  |
| **Region** | 498 |  | 0.4248 |  |  |  |  | 0.536 |
| East Midlands |  | 16.61 (12.29-21.96) |  | 1.52 (1.04-2.21) | 1.32 (0.88-1.97) | 1.10 (0.86-1.41) | 0.431 |  |
| East of England |  | 15.29 (11.67-19.68) |  | 1.40 (0.98-2.00) | 1.27 (0.87-1.86) | 0.97 (0.78-1.21) | 0.780 |  |
| London |  | 10.96 (8.36-14.10) |  | Ref | Ref | 1.01 (0.81-1.26) | 0.926 |  |
| North East |  | 14.71 (9.32-22.07) |  | 1.34 (0.83-2.17) | 1.13 (0.67-1.92) | 0.69 (0.47-1.01) | 0.058 |  |
| North West |  | 13.85 (10.66-17.68) |  | 1.26 (0.89-1.79) | 1.13 (0.77-1.65) | 1.14 (0.92-1.41) | 0.235 |  |
| South East |  | 14.38 (11.45-17.83) |  | 1.31 (0.94-1.83) | 1.16 (0.81-1.66) | 1.08 (0.90-1.31) | 0.395 |  |
| South West |  | 17.21 (13.00-22.35) |  | 1.57 (1.09-2.26) | 1.36 (0.92-2.01) | 1.09 (0.87-1.37) | 0.458 |  |
| West Midlands |  | 14.88 (11.27-19.28) |  | 1.36 (0.95-1.95) | 1.29 (0.88-1.88) | 1.06 (0.85-1.33) | 0.616 |  |
| Yorkshire and Humber |  | 13.54 (0.92-18.07) |  | 1.24 (0.84-1.82) | 1.11 (0.73-1.67) | 0.94 (0.73-1.21) | 0.617 |  |

Values are Rates (95% CI) per 1,000,000 children or Incident Rate Ratio (IRR) (95% CI).
\*Population used is children aged 5-17 years

Overall, there was little evidence that risk changed over the 4 year period (IRR 1.03 (0.96-1.11), p_trend_=0.863), or that the underlying trend was different by sex (p_trend_=0.863), ethnicity (p_trend_=0.174), age (p_trend_=0.106), deprivation (p_trend_=0.437), or region of England (p_trend_=0.536). However, there was some evidence that deaths by likely suicide in Asian or Asian British CYP period (IRR 1.48 (1.08- 2.02), p_trend_=0.015), and those in the least deprived quintiles (IRR 1.20 (1.01-1.42), p_trend_=0.041) may be increasing over the study period. There was weaker evidence for an increase in the risk for the youngest age group (5-15 years) (IRR 1.11 (0.99-1.24), p_trend_=0.084), and a decrease in risk for CYP in the North East of England (IRR 0.69 (0.47-1.01), p_trend_=0.058).

Finally, there was little evidence that sex, age or regional associations with suicide risk (reported in Table 4) were different across ethnic groups (Table 5), in the unadjusted (Sex, p=0.5055; Age p=0.8699; Region, p=0.7751) or adjusted analysis (Sex, p=0.4962; Age p=0.8772; Region, p=0.7426). There was however strong evidence that the relationship between deprivation and suicide risk was different between ethnic groups in both models (both p<0.001), with decreasing deprivation associated with increasing risk of suicide in white children (Unadjusted IRR 1.08 (1.00-1.16); Adjusted, IRR 1.12 (1.03-1.21)), and decreasing risk in CYP of Asian (Unadjusted IRR 0.56 (0.45- 0.71); Adjusted, IRR 0.52 (0.41-0.65)), Black (Unadjusted IRR 0.45 (0.24-0.48); Adjusted, IRR 0.31 (0.21-0.44)) and Mixed/Other (Unadjusted IRR 0.76 (0.63-0.92); Adjusted, IRR 0.73 (0.60-0.89) backgrounds). The observed (unadjusted) risk for each group, by quintiles of deprivation, is shown in Figure 1.

**Figure 1.**
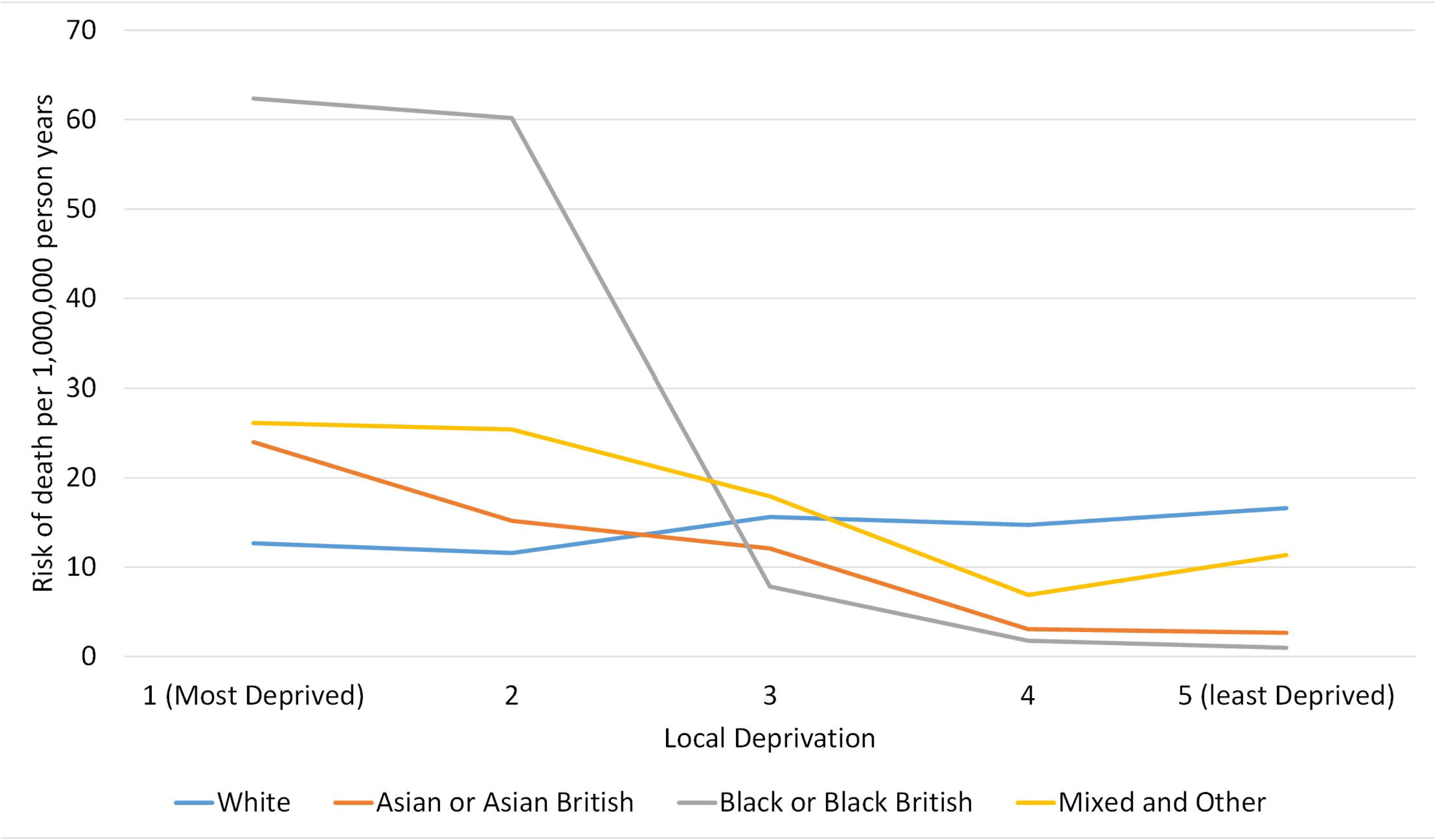
Rate of Suicide per 1,000,000 person years (5-17 year olds), by local deprivation; split by ethnic group.

**Table 5.** Relative risks of suicide by child characteristics, split by ethic group.

| Characteristic | n | White | Asian | Black | Mixed and Other | p |
| --- | --- | --- | --- | --- | --- | --- |
| Unadjusted Model |  |  |  |  |  |  |
| Sex | 497 |  |  |  |  |  |
| Female |  | Ref | Ref | Ref | Ref | 0.5055 |
| Male |  | 1.67 (1.35-2.07) | 1.91 (0.93-3.94) | 0.97 (0.41-2.34) | 1.26 (0.72-2.19) |  |
| Age (Years) | 498 |  |  |  |  |  |
| 5-15 |  | Ref | Ref | Ref | Ref | 0.8699 |
| 16/17 |  | 7.19 (5.84-8.84) | 7.88 (3.95-15.72) | 5.26 (2.19-12.65) | 6.31 (3.64-10.93) |  |
| Decreasing Deprivation Quintile | 493 | 1.08 (1.00-1.16) | 0.56 (0.45-0.71) | 0.34 (0.24-0.48) | 0.76 (0.63-0.92) | <0.001 |
| Region | 498 |  |  |  |  |  |
| East Midlands |  | 1.01 (0.62-1.64) | 2.26 (0.68-7.49) | 1.16 (0.14-9.23) | 3.21 (1.22-8.44) | 0.7751 |
| East of England |  | 1.17 (0.75-1.82) | 1.48 (0.39-5.58) | 1.58 (0.34-7.44) | 0.93 (0.26-3.39) |  |
| London |  | Ref | Ref | Ref | Ref |  |
| North East |  | 0.92 (0.58-1.65) | 2.09 (0.26-16.70) | NA | 1.46 (0.19-11.40) |  |
| North West |  | 0.92 (0.58-1.45) | 1.12 (0.34-3.73) | 1.46 (0.31-6.89) | 1.88 (0.68-5.18) |  |
| South East |  | 0.99 (0.65-1.52) | 0.96 (0.25-3.61) | 2.60 (0.78-8.62) | 1.92 (0.78-4.73) |  |
| South West |  | 1.28 (0.82-2.00) | NA | NA | 1.06 (0.23-4.85) |  |
| West Midlands |  | 1.05 (0.65-1.69) | 1.15 (0.38-3.52) | 1.55 (0.41-5.84) | 2.41 (0.98-5.92) |  |
| Yorkshire and Humber |  | 0.90 (0.55-1.48) | 1.77 (0.58-5.42) | NA | 1.69 (0.52-5.39) |  |
| Adjusted Model* |  |  |  |  |  |  |
| Sex | 497 |  |  |  |  |  |
| Female |  | Ref | Ref | Ref | Ref | 0.4962 |
| Male |  | 1.67 (1.35-2.06) | 1.88 (0.91-3.88) | 0.94 (0.39-2.26) | 1.25 (0.72-2.17) |  |
| Age (Years) | 498 |  |  |  |  |  |
| 5-15 |  | Ref | Ref | Ref | Ref | 0.8772 |
| 16/17 |  | 7.22 (5.88-8.88) | 7.98 (4.00-15.92) | 5.23 (2.17-12.57) | 6.35 (3.67-11.00) |  |
| Decreasing Deprivation Quintile | 493 | 1.12 (1.03-1.21) | 0.52 (0.41-0.65) | 0.31 (0.21-0.44) | 0.73 (0.60-0.89) | <0.001 |
| Region | 498 |  |  |  |  |  |
| East Midlands |  | 1.01 (0.62-1.65) | 1.90 (0.57-6.34) | 0.65 (0.08-5.28) | 2.96 (1.12-7.81) | 0.7426 |
| East of England |  | 1.21 (0.77-1.89) | 0.98 (0.26-3.75) | 0.49 (0.10-2.34) | 0.75 (0.21-2.76) |  |
| London |  | Ref | Ref | Ref | Ref |  |
| North East |  | 0.86 (0.48-1.53) | 2.07 (0.26-16.59) | NA | 1.57 (0.20-12.29) |  |
| North West |  | 0.87 (0.83-1.37) | 1.56 (0.47-5.17) | 2.39 (0.51-11.28) | 2.09 (0.76-5.75) |  |
| South East |  | 1.05 (0.68-1.62) | 0.51 (0.13-1.96) | 0.69 (0.20-2.36) | 1.39 (0.55-3.51) |  |
| South West |  | 1.30 (0.83-2.03) | NA | NA | 0.89 (0.19-4.08) |  |
| West Midlands |  | 1.00 (0.62-1.62) | 1.55 (0.51-4.75) | 2.07 (0.55-7.80) | 2.69 (1.09-6.62) |  |
| Yorkshire and Humber |  | 0.86 (0.53-1.41) | 2.77 (0.90-8.52) | NA | 1.82 (0.57-5.80) |  |

Values are Incident Rate Ratio (IRR) (95% CI) compared to reference categories.
\* Adjusted for other measures (sex, age, region and deprivation)

## DISCUSSION

### Key Findings

During a four year period, between April 2019 and March 2023, a total of 498 CYP likely died of suicide; and while there was some rise during the first, and second year of the pandemic, risks afterwards appear similar to a year before COVID, and the lockdown(9). Risks were substantially higher in older children and males. However, the characteristics appeared similar across the time period, despite the broad social changes that have occurred, with similar sex, ethnicity, age and geographic locations across the cohort.

There was also a likely increase in the number of children identifying as LGBT+ across the 4 years, and this was highest in the 3^rd^ year of the cohort (the second year of the pandemic). However, in the most recent data, the proportion appears to have dropped to similar levels to 2019-20. Hanging was the most common method of death, and previous self-harm the most prevalent previous diagnosis; with no real change seen over the 4 year time period. Equally, an acute hospital admission with self-harm was associated with a 8-9 fold increase in the next 2-28 days in the risk of death, compared to other periods of the child’s life. Acute hospital admissions with ICD coded anxiety or mood disorders were also associated with a higher subsequent risk, but due to small numbers, statistical precision was poor. Overall risk of suicide differed by ethnicity and measures of local deprivation; but the relationship was complicated, with increasing risk, as deprivation reduced, in children in white backgrounds, and decreasing risk in other groups; with similar risks seen in the middle quintile of deprivation.

### Limitations

This work is based on the ongoing national data collection from the NCMD, and previous work has shown good validation and coverage (23), although there remain several limitations to our analysis. While we had some missing data on some demographics (e.g. ethnicity), data completeness was generally good, and the primary analysis of risks was based on the statutory reporting of deaths; although we did only have HES data on the first 3 years of the cohort, and so changes across the last year are not included in this work. In addition, this data was limited to acute-healthcare delivery (rather than wider mental health services) and therefore may not be an accurate reflection of the prevalence of mental health disorders for all CYP who had subsequently died by suicide (24). In addition, categorisation of each death (including coding of LGBT+ and method of death) may have been based on initial data, with some deaths in this analysis waiting full CDOP review; although previous NCMD work has suggested good validity with this methodology(7). Finally, child suicides in England are fortunately rare, which limits the precision of our estimates, and hence their interpretation.

### In Context

Our results offer a valuable insight into both the stability and shifting patterns of risk amid significant societal disruptions, such as the COVID-19 pandemic. Previous research has highlighted concerns about the mental health impact of the pandemic on young people, including increased rates of anxiety, depression, and self-harm(25, 26). While our findings show a modest, but not significant, increase in suicides during the early pandemic years, the return to pre-pandemic levels by 2022–23 contrasts with fears of a sustained surge in youth suicide rates, aligning with more nuanced international studies that report variable effects across age groups and contexts(27, 28). Equally the consistently higher suicide risk in older children and males is well supported by longstanding epidemiological patterns(29), while the emergence of increased risk in children from the most deprived quintile, in some ethnic groups, echoes concerns about widening health inequalities post-pandemic(30). The complex interaction between ethnicity, deprivation, and suicide risk is particularly notable and adds additional depth to the existing literature, which often lacks sufficient granularity to explore these intersections meaningfully. The overall, increase in children identifying as LGBT+ in the cohort also aligns with growing recognition of this group’s heightened vulnerability to mental health needs and suicide risk(31, 32). The overall evidence on suicide risk in children and young people with gender dysphoria is poor, although these CYP often experience many of the factors known to be associated with increased risk including experiences of bullying, poor mental health and autism (33). The Cass review(19), found no strong evidence that gender-affirming medical treatments reduce suicide risk. The overall prevalence (identified from acute-healthcare linkage) of ASD was around 5% in this work, and while this has been recognised as a risk factor for suicide(34), we saw little evidence of any change in the prevalence across the study.

The strong temporal associations with prior self-harm further reinforce known clinical risk markers(35). This work highlights opportunities for professionals, including mental health services to intervene following an episode of self-harm, which has been highlighted in previous national reports(36, 37). Recent national guidance for practitioners working with CYP and adults, in contact with mental health services has also been made available to improve clinical practice (38).

## Conclusion

In England, suicide rates do not appear to be increasing, and the methods of suicide remain static. However, the role of deprivation and suicide risk appears to be different between children of different ethnic groups. While acute-hospital admission with a range of mental health disorders do not appear to predict likely suicide in the subsequent month, there was a strong association with self-harm events.

## Author Contributions

DO: I declare that I contributed to writing (original draft and review and editing), conceptualization, data curation, formal analysis, funding acquisition, investigation, methodology, project administration and resources; and that I have seen and approved the final version.

DK: I declare that I contributed to writing (original draft and review and editing), conceptualization, data curation and investigation; and that I have seen and approved the final version.

TW: I declare that I contributed to writing (original draft and review and editing), conceptualization, data curation, funding acquisition, investigation, methodology, project administration and resources; and that I have seen and approved the final version.

SS: I declare that I contributed to writing (original draft and review and editing), conceptualization, data curation, funding acquisition, investigation, methodology, project administration and resources; and that I have seen and approved the final version.

PC: I declare that I contributed to writing (original draft and review and editing), conceptualization, data curation and investigation; and that I have seen and approved the final version.

KL: I declare that I contributed to writing (original draft and review and editing), conceptualization, data curation, funding acquisition, investigation, methodology, project administration and resources; and that I have seen and approved the final version.

## Acknowledgments

We thank all Child Death Overview Panels (CDOPs) who submitted data for the purposes of this report and all child death review professionals for submitting data and providing additional information when requested.

Parent and public involvement is at the heart of the NCMD programme. We are indebted to Charlotte Bevan (Sands - Stillbirth and Neonatal Death Charity), Ann Chalmers (Child Bereavement UK) and Jenny Ward (Lullaby Trust), who represent bereaved families on the NCMD programme steering group.

We thank the NCMD team for technical and administrative support.

David Odd had full access to all the data in the study and takes responsibility for the integrity of the data and the accuracy of the data analysis.

## Data Availability Statement

Aggregate data may be available on request to the corresponding author, and subject to approval by HQIP.

## Ethics approval and consent to participate

The NCMD legal basis to collect confidential and personal level data under the Common Law Duty of Confidentiality has been established through the Children Act 2004 Sections M - N, Working Together to Safeguard Children 2018 (https://consult.education.gov.uk/child-protection-safeguarding-and-family-law/working-together-to-safeguard-children-revisions-t/supporting_documents/Working%20Together%20to%20Safeguard%20Children.pdf) and associated Child Death Review Statutory & Operational Guidance https://assets.publishing.service.gov.uk/government/uploads/system/uploads/attachment_data/file/859302/child-death-review-statutory-and-operational-guidance-england.pdf). The NCMD legal basis to collect personal data under the General Data Protection Regulation (GDPR) without consent is defined by GDPR Article 6 (e) Public task and 9 (h) Health or social care (with a basis in law).

